# Voluntary visual imagery and non-suicidal self-injury in patients with emotionally unstable personality disorder

**DOI:** 10.64898/2026.06.23.26356159

**Authors:** Jenya Shaji, Ratheesh S R, Rajith Ravindren

## Abstract

**Background:** Emotionally Unstable Personality Disorder (EUPD) is characterised by emotional dysregulation, unstable self-identity, interpersonal difficulties, dissociation, and a high prevalence of non-suicidal self-injury (NSSI). Mental imagery plays a role in emotion processing and autobiographical memory. Yet little is known about the relation between voluntary visual imagery and self-harm in EUPD. The study aimed to examine the imagery characteristics, including visual imagery vividness and synaesthetic-like experiences, and their association with NSSI in EUPD.

**Method:** Forty adults aged 18-45 years meeting ICD-10 Diagnostic Criteria for Research (ICD-10 DCR) for EUPD were recruited through purposive sampling. Visual imagery was assessed using the Vividness of Visual Imagery Questionnaire-2 (VVIQ-2). NSSI and its functions were assessed using the Inventory of Statements About Self-Injury (ISAS). Synaesthesia-like experiences were screened using a seven-item questionnaire developed for the study. Group comparisons were performed using independent-samples t-tests, and correlations were assessed using Pearson’s correlation coefficient.

**Results:** Twenty-nine patients (72.5%) with EUPD had NSSI. Participants with NSSI had significantly higher mean VVIQ-2 scores than those without NSSI (118.48 +/-24.81 vs. 93.36 +/-35.66; p = 0.016; Cohen’s d = 0.89). VVIQ-2 scores correlated positively with intrapersonal ISAS functions (r = 0.38, p = 0.015) but not interpersonal functions (r = 0.19, p = 0.22). Three participants (7.5%) demonstrated imagery scores compatible with probable hyperphantasia. Four participants reported synaesthesia-like experiences.

**Conclusions:** Greater visual imagery was associated with non-suicidal self-harm in patients with EUPD. Imagery vividness was predominantly associated with intrapersonal functions of non-suicidal self-harm, particularly affect regulation, rather than interpersonal motivations. These findings suggest that visual imagery may represent a previously under-recognized cognitive factor contributing to emotional dysregulation and self-injurious behaviour in EUPD. Assessment of imagery characteristics may have clinical relevance when designing psychotherapeutic interventions for EUPD.

## Introduction

Emotionally unstable personality disorder (EUPD) is characterised by a pervasive pattern of emotional dysregulation, self-identity issues, and interpersonal problems.^1^ EUPD in adults has a point prevalence of 5.9%.^2^ It has an even higher prevalence in psychiatric outpatient services and hospital settings.^3^ EUPD is associated with substantial psychosocial impairment, including reduced educational and occupational achievement, unstable interpersonal relationships, greater partner conflict, increased sexual risk-taking, limited social support, and lower life satisfaction. Anxiety disorders, mood disorders, PTSD, and substance use are more common in this group.^4^ The patients are considered to be difficult to deal with by clinicians due to their demanding behaviour and poor adherence to treatment. This group of patients causes significant strain to the mental health services worldwide.^3^

The underlying pathology of this disorder remains unclear. Studies have found that it has an estimated heritability of 40–50%.^5^ Increased amygdala activation and reduced frontal activity are consistently reported in patients. Neurophysiological studies and self-reports have found that patients have low pain sensitivity and frequently have dissociative symptoms like depersonalisation, derealisation, and numbing. In addition to a high risk of suicide, non-suicidal self-harm is also seen more in EUPD. Non-suicidal self-harm is taken to reduce stress and terminate dissociation. This has been validated by a study that reports reduced amygdala activation in EUPD on self-harm.^6^

Patients often exhibit poor social cognition and social information processing skills, which can lead to difficulties in interpersonal relationships.^7^ They also have a lack of coherence in thoughts and feelings, which may be the reason for unstable self-identity and emotional dysregulation.^8^ The underlying brain changes causing these are still not clear.

Mental images play an important role in emotion processing, autobiographic memory, and social cognition. The ability to have mental images lies on a spectrum. Some people cannot have voluntary images -a condition called aphantasia while others can have extremely vivid images,called hyperphantasia.^9^ Most people have visual images in between. People with more visual imagery are also likely to have a condition called synaesthesia, where stimulation of one sensory modality causes unusual experiences in another unrelated modality.^10,11^

Variations in mental imagery may cause emotional dysregulation and psychiatric morbidities. Absence of visual imagery was found to negatively impact the ability to simulate the past and future.^12^ People with hyperphantasia were found to have greater autobiographical memory and imagination.^13^ Fewer intrusive images were reported in a study that used the trauma film paradigm as a laboratory model of PTSD in aphantasia.^14^Reduced somatic responses in the form of poor GSR responses in emotional conditions were also found in aphantasia, showing that reduced imagery is protective against emotional disorders.^15^ As a corollary, increased imagery may be associated with intrusive images, increasing the risk of PTSD and other emotional problems.

Few studies have examined what happens to imagery, especially voluntary visual imagery, and its variants in EUPD, and whether the variation in visual imagery plays a role in the symptoms seen in EUPD. Understanding visual imagery may help in unravelling novel insights into the mechanisms of emotional dysregulation, unstable self-identity, and self-harm in EUPD. The study aimed to study visual imagery characteristics, including imagery vividness and synesthetic experiences in EUPD, and their relation to non-suicidal self-harm.

## Method

Forty adults (18–45 years) meeting ICD -10 Diagnostic Criteria for Research (ICD-10 DCR) criteria for EUPD were included via purposive sampling.^16^ Those with intellectual disability, sensory impairment, or comorbid depressive or other major psychiatric disorders were excluded from the study. After obtaining informed consent from the patients, the following tools were administered.

### Data collection tools

#### 1. Vividness of Visual Imagery Questionnaire-2 (VVIQ-2)^17^

The VVIQ - 2 typically asks participants to visualise 32 scenarios which they rate on a five-point scale from 1 (‘no image at all, you only know that you are thinking of the object’) to 5 (‘perfectly clear and vivid as real seeing’). The questionnaire has demonstrated high internal consistency reliability (Cronbach’s alpha ≈ 0.91) and good construct validity in measuring visual imagery vividness.^18^ VVIQ-2 scores were categorized operationally into probable aphantasia (32–39), very weak imagery (40–63), low imagery (64–94), average imagery (95– 127), high imagery (128–149), and very vivid imagery/probable hyperphantasia (150–160), based on score distribution and prior literature on imagery vividness extremes.^13^

#### 2. Inventory Of Statement About Self Injury (ISAS) ^19^

The first section of the ISAS assesses the lifetime frequency of the twelve Non-Suicidal Self Injury (NSSI) behaviours which are performed “intentionally (i.e., on purpose) and without suicidal intent.” The behaviors assessed include banging/hitting oneself, biting, burning, carving, cutting, wound picking, needle-sticking, pinching, hair pulling, rubbing skin against rough surfaces, severe scratching, and swallowing chemicals.

Those having one or more NSSI behaviors are instructed to complete the second section of the ISAS. The second section assesses 13 potential functions of NSSI: affect regulation, anti-dissociation, anti-suicide, autonomy, interpersonal boundaries, interpersonal influence, marking distress, peer bonding, self-care, self-punishment, revenge, sensation seeking, and toughness. Each function is assessed by three items, rated as “0-not relevant,” “1-somewhat relevant,” or “2-very relevant” to the individual’s “experience of non-suicidal self-harm”. Scores for each of the 13 ISAS functions can range from 0 to 6. ISAS functions exhibited a two-factor structure. Interpersonal, by which NSSI is socially reinforced, and intrapersonal, by which NSSI is self-reinforced. The first factor represented interpersonal functions - autonomy, interpersonal boundaries, interpersonal influence, peer-bonding, revenge, sensation seeking, and toughness. The second factor represented intrapersonal functions-affect-regulation, anti-dissociation, anti-suicide, marking distress, self-care, and self-punishment. Coefficient alphas for the interpersonal and intrapersonal scales were 0.88 and 0.80, respectively, indicating excellent internal consistency.

#### 3. Questionnaire to screen synaesthesia

Synesthetic experiences were assessed using a brief seven-item questionnaire developed for the study. The questionnaire screened for common forms of synaesthesia and related cross-modal perceptual experiences. ^10,20^ Participants were asked whether they experienced:

1. colour associations with letters or numbers (grapheme-colour synaesthesia)
2. colour associations with weekdays or months,
3. spatial locations associated with temporal sequences such as weekdays, months, or years (spatial-sequence synaesthesia)
4. colour experiences triggered by sounds (sound-colour synaesthesia)
5. taste sensations evoked by words (lexical-gustatory synaesthesia)
6. tactile sensations elicited by smells (olfactory-tactile synaesthesia)
7. any other unusual automatic blending of sensory experiences not covered by the preceding questions.

Each item was presented with examples to facilitate understanding. Responses were recorded in a dichotomous format (Yes/No).

### Data Analysis

The data were analysed using (JASP) (Version 0.19.3.0).^21^ Descriptive statistics (mean and standard deviation) were calculated to summarize the distribution of data. The assumption of normality was assessed using the Shapiro -Wilk test. Levene’s test was used to assess the equal variance criterion. An independent sample t-test was used to find the difference in sample means. Pearson’s correlation coefficient was used to find the correlation between imagery score and the ISAS scores. For all statistical tests, the significance level was predetermined at 0.05. All data were included in the analysis, and no missing data were reported.

## Results

Among the 40 patients, 36 were female, and 4 were male. The mean age of the group was 25.2 years. The mean male age was 27.8 years, while the mean female age was 24.9 years.

Self-harm was reported by 29 patients (72.5%). The mean age of those with self-harm was 24.5 years, while the mean age of those with no self-harm was 27.3 years.

Among the four males, 3 had self-harm, while among the 36 females, 26 had reported self-harm. The prevalence of self-harm did not differ significantly by sex (p=0.9). (**Table 1**)

**Table 1.** Sociodemographic and clinical characteristics (N = 40)

| Variable | Value |
| --- | --- |
| Age, mean (SD), years | 25.2 (6.6) |
| Female, n (%) | 36 (90%) |
| Male, n (%) | 4 (10%) |
| Female age, mean (SD), years | 24.9 (6.7) |
| Male age, mean (SD), years | 27.8 (5.7) |
| Self-harm present, n (%) | 29 (72.5%) |
| No self-harm, n (%) | 11 (27.5%) |
| **Types of non-suicidal self-harm** |  |
| Cutting | 23 (57.5%) |
| Hitting self | 16 (40%) |
| Pulling hair | 14 (35%) |
| Severe scratching | 11 (27.5%) |
| Biting | 10 (25%) |
| Pinching | 8 (20%) |
| Burning | 5 (12.5%) |
| Swallowing dangerous substances | 5 (12.5%) |
| Rubbing skin against rough surfaces | 4 (10%) |

The common reasons given for non-suicidal self-injury were affect regulation and self-punishment (**Table 2**)

**Table 2:** Reasons for NSSI among patients (N=29)

| Reasons | Mean Score(maximum=6) |
| --- | --- |
| Affect regulation | 4.4 |
| Self-punishment | 4.2 |
| Marking distress | 3.2 |
| Anti-dissociation | 3.1 |
| Anti-suicide | 3.1 |
| Toughness | 2.5 |
| Interpersonal boundaries | 2.2 |
| Revenge | 2.0 |
| Interpersonal influence | 1.9 |
| Self-care | 1.9 |
| Autonomy | 1.9 |
| Sensation seeking | 1.0 |
| Peer-bonding | 0.97 |

VVIQ-2 was compared between those with self-harm and those without. The Shapiro-Wilk test and Levene’s test of equality of variance were not significant. VVIQ-2 scores were significantly higher among participants with NSSI (mean = 118.48, standard deviation = 24.81) than among those without NSSI (mean = 93.36, standard deviation = 35.66), p = 0.016, Cohen’s d = 0.89

VVIQ2 did not differ between males and females (mean male score=103.5, mean female score=112.5, p=0.6)

VVIQ-2 scores correlated positively with intrapersonal ISAS functions (r = 0.38, p = 0.015) but not with interpersonal functions (r=0.19, p=0.22) (**Table 3**)

**Table 3.** Association of VVIQ-2 scores with self-harm and ISAS functional domains.

| Variable | Mean (SD) / r | P value | Effect size |
| --- | --- | --- | --- |
| Self-harm present(n=29) | 118.48 (24.81) | **0.016** | Cohen's $d = 0.89$ |
| No self-harm(n=11) | 93.36 (35.66) |  |  |
| ISAS intrapersonal functions | $r = 0.38$ | **0.015** | — |
| ISAS interpersonal functions | $r = 0.19$ | 0.22 | — |

Three patients (7.5%) demonstrated very high imagery vividness (VVIQ-2 >150), while ten (25%) had high visual imagery in the range 128–149. None had a very low score of less than 40. Four patients reported synaesthesia-like experiences, including grapheme-colour associations (n=2), colour associations with temporal sequences (n=1), and spatial representations of time (n=1). (**Table 4**)

**Table 4.** VVIQ-2 Imagery categories and synaesthesia characteristics.

| Variable | Category/Type | n (%) |
| --- | --- | --- |
| **VVIQ2 imagery category** | Probable aphantasia (32–39) | 0 |
|  | Very weak imagery (40–63) | 3 (7.5%) |
|  | Low imagery (64–94) | 9 (22.5%) |
|  | Average imagery (95–127) | 15 (37.5%) |
|  | High imagery (128–149) | 10 (25%) |
|  | Probable hyperphantasia (150–160) | 3 (7.5%) |
| **Synaesthesia-like experiences** | Grapheme–colour | 2 (5%) |
|  | Days/months–colour associations | 1 (2.5%) |
|  | Spatial arrangement of time | 1 (2.5%) |

## Discussion

To our knowledge, this is among the first studies to investigate imagery vividness in relation to self-harm behaviour in EUPD.

NSSI is defined as self-directed and intentional behavior that causes harm or destruction to bodily tissue without the intent to die.^22^ The common non -suicidal self -harm reported by the study subjects included cutting skin, hitting, pulling hair, severe scratching, biting, and pinching. NSSI was seen in three-quarters of the sample, with no gender specific differences. Similar rates were reported previously.^3^

Affect regulation, self-punishment, marking distress, anti-suicide, and anti-dissociation were the common reasons given for NSSI. All these reasons indicate intrapersonal functions of the NSSI. These findings suggest that self-harm in EUPD occurs mostly due to internal factors-mainly emotional dysregulation-rather than for secondary gains. Neurophysiological and fMRI studies have reached a similar conclusion. Patients with EUPD have pain hyposensitivity, overactive amygdala, and reduced parasympathetic tone. Self-injury reduces amygdala hyperactivity and increases functional connectivity between the amygdala and the superior frontal gyrus. NSSI thus helps in correcting the disturbed emotional circuits in EUPD.^6^

Three patients (7.5%) demonstrated very high imagery vividness (VVIQ-2 >150), consistent with probable hyperphantasia. This proportion appears higher than the prevalence of 3% in the community samples.^9^ But in the context of the small sample size and absence of a control group, this finding should be considered exploratory. Another 25% had high visual imagery in the range 128–149. None had a very low score of less than 40 - scores which are reported by people with aphantasia.

Patients with non-suicidal self-harm showed significantly higher imagery vividness. The observed effect size was large (Cohen’s d = 0.89), suggesting that imagery vividness may have clinically meaningful associations with self-harm behaviour. Higher imagery vividness was associated with greater endorsement of intrapersonal functions of NSSI, particularly those related to emotional regulation and distress reduction.

Those with stronger visual imagery may experience more vivid autobiographical memories and may be involved in more affect-laden simulations of the future. This may potentially increase emotional dysregulation and dissociative symptoms in patients. They may, in turn, resort to more self-harm behaviours to calm themselves down. Such mechanisms may explain our finding of increased self-harm being associated with more vivid imagery. The observation is consistent with reports that found individuals with aphantasia have less intrusive images and emotional dysregulation. ^14,15^

Together, these raise the possibility that excessive visual imagery may contribute to self-harm behaviour in EUPD. The exact mechanism of how this happens can be revealed only by in-depth studies using a larger sample size and functional brain imaging.

Synesthesia-like experiences were reported by four patients. Given the small number of cases and the absence of a validated synaesthesia assessment, these findings should be considered preliminary,

The study is limited by its small sample size. An estimation of hyperphantasia or synaesthesia from this sample will be considered only as exploratory. Besides, we have not used a standard synaesthesia questionnaire for this study. The absence of a healthy control group also limits the conclusions regarding whether imagery vividness is elevated relative to the general population. The direction of the association between imagery vividness and non-suicidal self-injury cannot be ascertained by the study design.

Greater visual imagery was associated with non-suicidal self-harm in patients with EUPD. Imagery vividness was predominantly associated with intrapersonal functions of non-suicidal self-harm, particularly affect regulation, rather than interpersonal motivations. Visual imagery may represent a previously under-recognized cognitive factor contributing to emotional dysregulation and self-injurious behaviour in EUPD. Assessment of imagery characteristics may be clinically relevant as many of the psychotherapeutic interventions employ imagery-based techniques.

## Data Availability

All data produced in the present study are available upon reasonable request to the authors

## Notes

### Competing Interest Statement

The authors have declared no competing interest.

### Author Declarations

Institute of Mental Health and Neurosciences(IMHANS), Kozhikode Ethics Committee (Reg No: EC/NEW/INST/2023/4210) Approval No: IMHANS/IEC/CPT2024/010 dated 21/08/2024

## Reference

1. American Psychiatric Association. Diagnostic and statistical manual of mental disorders. 5th ed. Washington (DC): American Psychiatric Publishing; 2013.

2. Grant BF, Chou SP, Goldstein RB, Huang B, Stinson FS, Saha TD, Smith SM, Dawson DA, Pulay AJ, Pickering RP, Ruan WJ. Prevalence, correlates, disability, and comorbidity of DSM-IV borderline personality disorder: results from the Wave 2 National Epidemiologic Survey on Alcohol and Related Conditions. Journal of clinical psychiatry. 2008 Apr 23;69(4):533

3. Gunderson JG, Herpertz SC, Skodol AE, Torgersen S, Zanarini MC. Borderline personality disorder. Nature reviews disease primers. 2018 May 24;4(1):18029.

4. Bohus M, Stoffers-Winterling J, Sharp C, Krause-Utz A, Schmahl C, Lieb K. Borderline personality disorder. The lancet. 2021 Oct 23;398(10310):1528–40.

5. Distel MA, Trull TJ, Derom CA, Thiery EW, Grimmer MA, Martin NG, Willemsen G, Boomsma DI. Heritability of borderline personality disorder features is similar across three countries. Psychological medicine. 2008 Sep;38(9):1219–29.

6. Reitz, S., Kluetsch, R., Niedtfeld, I., Knorz, T., Lis, S., Paret, C., Kirsch, P., Meyer-Lindenberg, A., Treede, R.D., Baumgaertner, U. and Bohus, M., 2015. Incision and stress regulation in borderline personality disorder: neurobiological mechanisms of self-injurious behaviour. The British Journal of Psychiatry, 207(2), pp.165–172.

7. Nemeth N, Matrai P, Hegyi P, Czeh B, Czopf L, Hussain A, Pammer J, Szabo I, Solymar M, Kiss L, Hartmann P. Theory of mind disturbances in borderline personality disorder: A meta-analysis. Psychiatry research. 2018 Dec 1;270:143–53.

8. Luyten P, Campbell C, Allison E, Fonagy P. The mentalizing approach to psychopathology: State of the art and future directions. Annual review of clinical psychology. 2020 May 7;16(1):297–325.

9. Zeman A. Aphantasia and hyperphantasia: exploring imagery vividness extremes. Trends in cognitive sciences. 2024 May 1;28(5):467–80.

10. Ramachandran VS, Hubbard EM. Synaesthesia--a window into perception, thought and language. Journal of consciousness studies. 2001 Dec 1;8(12):3–4.

11. Barnett KJ, Newell FN. Synaesthesia is associated with enhanced, self-rated visual imagery. Consciousness and cognition. 2008 Sep 1;17(3):1032–9.

12. Dawes AJ, Keogh R, Robuck S, Pearson J. Memories with a blind mind: Remembering the past and imagining the future with aphantasia. Cognition. 2022 Oct 1;227:105192.

13. Milton, F., Fulford, J., Dance, C., Gaddum, J., Heuerman-Williamson, B., Jones, K., Knight, K.F., MacKisack, M., Winlove, C. and Zeman, A., 2021. Behavioral and neural signatures of visual imagery vividness extremes: Aphantasia versus hyperphantasia. Cerebral cortex communications, 2(2), p.tgab035.

14. Keogh R, Wicken M, Pearson J. Fewer intrusive memories in aphantasia: using the trauma film paradigm as a laboratory model of PTSD. PsyArXiv [Preprint]. 2023. Available from: doi:10.31234/osf.io/7zqfe

15. Zeman A, Milton F, Della Sala S, Dewar M, Frayling T, Gaddum J, Hattersley A, Heuerman-Williamson B, Jones K, MacKisack M, Winlove C. Phantasia–The psychological significance of lifelong visual imagery vividness extremes. Cortex. 2020 Sep 1;130:426–40

16. World Health Organization. The ICD-10 classification of mental and behavioural disorders: diagnostic criteria for research. Geneva: World Health Organization; 1993.

17. Marks DF. New directions for imagery research. J Ment Imagery. 1995;19:153–167.

18. Campos A. Internal consistency and construct validity of two versions of the Revised Vividness of Visual Imagery Questionnaire. Perceptual and Motor Skills. 2011 Oct;113(2):454–60.

19. Klonsky ED, Glenn CR. Assessing the functions of non-suicidal self-injury: Psychometric properties of the Inventory of Statements About Self-injury (ISAS). Journal of psychopathology and behavioral assessment. 2009 Sep;31(3):215–9.

20. Eagleman DM, Kagan AD, Nelson SS, Sagaram D, Sarma AK. A standardized test battery for the study of synesthesia. Journal of neuroscience methods. 2007 Jan 15;159(1):139–45.

21. JASP Team. JASP (Version 0.19.3.0) [Computer software]. Amsterdam: University of Amsterdam; 2025.

22. Brickman LJ, Ammerman BA, Look AE, Berman ME, McCloskey MS. The relationship between non-suicidal self-injury and borderline personality disorder symptoms in a college sample. Borderline personality disorder and emotion dysregulation. 2014 Aug 27;1(1):14.

